# A Breast Cancer Polygenic Risk Score Validation in 15,490 Brazilians using Exome Sequencing

**DOI:** 10.1101/2024.04.21.24306089

**Authors:** Flávia Eichemberger Rius, Rodrigo Santa Cruz Guindalini, Danilo Viana, Júlia Salomão, Laila Gallo, Renata Freitas, Cláudia Bertolacini, Lucas Taniguti, Danilo Imparato, Flávia Antunes, Gabriel Sousa, Renan Achjian, Eric Fukuyama, Cleandra Gregório, Iuri Ventura, Juliana Gomes, Nathália Taniguti, Simone Maistro, José Eduardo Krieger, Yonglan Zheng, Dezheng Huo, Olufunmilayo I. Olopade, Maria Aparecida Koike, David Schlesinger

## Abstract

**Background:** Brazil has a highly admixed population. Polygenic Risk Scores (PRS) have mostly been developed from European population studies, and their application to other populations is challenging. To assess the use of PRS for breast cancer (BC) risk in Brazil, we evaluated four PRSs in the Brazilian population.

**Methods:** We analyzed a Brazilian cohort composed of 6,206 women with a history of breast cancer and 8,878 unphenotyped adults as controls. Genomic variants were imputed from exomes and scores were calculated for all samples.

**Results:** After excluding individuals with known pathogenic or likely pathogenic variants in *BRCA1*, *BRCA2*, *PALB2*, *PTEN*, or *TP53* genes, and first-degree relatives of the probands, 5,598 cases and 8,767 controls remained. Four PRS models were compared, and PRS_3820_ from Mavaddat *et al.* 2019 study showed the best performance, with an Odds Ratio (OR) of 1.43 per standard deviation increase (p-value: < 0.001) and an OR of 1.88 (p-value: < 0.001) for the top decile. PRS_3820_ also performed well for different ancestry groups: East Asian majority (OR 1.59, p-value 0.004), Non-European majority (OR 1.45, p-value: <0.001), and European majority (OR: 1.43, p-value: <0.001).

**Conclusion:** Among different PRS, the PRS_3820_ performed better in the highly admixed Brazilian population. This will allow a more precise BC risk assessment of mutation-negative women in Brazil.

## Introduction

Breast cancer (BC) is a critical global health concern, representing the most common cancer diagnosed among women^1^. In Brazil, over 70,000 women are diagnosed with BC every year, accounting for 30% of all cancers in the female population^2^.

Approximately 10% of all BC cases are attributable to germline pathogenic variants in susceptibility genes^3^. Rare variants in these genes account for approximately 25% of the genetic risk. The remaining genetic risk (∼75%) is derived from common, low penetrance variants that individually confer small risk, but which combined effect can be substantial^4–6^.

Genome-wide association studies (GWASs) have been predominantly carried out in European populations^7–10^. Evaluation of PRS across different genetic and environmental backgrounds is essential to enable the implementation of genetic risk stratification strategies for individuals from non-European populations^11^.

The Brazilian population exhibits a unique, highly admixed, genetic composition. It is mostly derived from a combination of Native Americans, Southern Europeans (Portuguese, Spanish, and Italian) that immigrated in the period 1500-1900, and Sub-Saharan Africans brought through extensive slave trading until the 1800s. More recently, from 1822 to the first half of the 1900s, other smaller waves of immigration also contributed to Brazil’s remarkable diversity, including Japanese, Lebanese, German, and Eastern Europeans^12^. Three in every four Brazilians have multiple genetic ancestries^13,14^. Given Brazil’s genetic diversity, any PRS developed in predominantly European populations requires validation before it can be used in clinical settings.

Several laboratory methods are available for genotyping variants directly or indirectly (imputation), including microarrays, whole exome sequencing (WES), and whole genome sequencing (WGS). WES offers an affordable and scalable alternative to arrays and WGS, while allowing for simultaneous rare and common variant genotyping. In this study, we evaluate four BC PRSs^7,8,15^ developed using WES in 15,490 Brazilians.

## Methods

### Study population

A total of 15,490 individuals were selected for this study, including 6,362 women with breast cancer history, and 9,128 adult unphenotyped controls. Both clinical and genetic data were collected from a database of a College of American Pathology (CAP)-accredited laboratory (Mendelics, São Paulo, SP, Brazil). All BC and control subjects provided Informed Consent for use of retrospective anonymized data for research purposes. Samples were anonymized before analysis. Clinical records such as BC histological type and age of diagnosis were obtained from genetic test requisitions. The study was IRB-approved (CAAE: 70112423.3.0000.0068).

### Exome sequencing and imputation

Exome sequencing data were generated from buccal swab or venous blood samples with standard protocol for Illumina Flex Exome Prep, using a custom probe set from Twist Biosciences. Sequencing was conducted in Illumina sequencers and the bioinformatics pipeline for data analysis followed Broad Institute’s GATK best practices (https://gatk.broadinstitute.org/hc/en-us/sections/360007226651-Best-Practices-Workflo ws), with alignment to GRCh38.

Imputation of exomes was based on a panel of 2,504 individuals of all ancestries from the 1000 Genomes Project (1KGP)^16^ on GRCh38 (2017 release) (https://www.internationalgenome.org/data-portal/data-collection/grch38). All regions captured from the exome sequencing comprehending at least 1x coverage, as well as off-target regions, were considered for the imputation, performed using Glimpse (v1.1.0) software^17^.

### Polygenic Risk Score Calculation

Four BC PRSs with publicly available summary statistics, from three different studies, were evaluated in this work: Khera *et al.* 2018^7^, with 5,218 variants (PGS Catalog ^18,19^ ID: PGS000015); Mavaddat *et al.* 2019^8^ PRSs (with 313 and 3,820 variants, respective PGS Catalog IDs: PGS000004 and PGS000007); and UK Biobank^15^ (UKBB) PRS obtained from a variant thresholding (p-value < 10e-5) on summary statistics for phenotype code 20001_1002, with 7,538 variants.

To overcome the constraints associated with variants derived from imputation, we filtered PRS variants based on their distance from exome bed kit and minor allele frequency (MAF). Variants with null betas (beta = 0) were removed from all PRSs. Additionally, the PRS from Mavaddat study, originally with 3,820 variants, had a pathogenic variant of moderate-penetrance in *CHEK2* gene (*CHEK2* p.Ile157Thr - Clinvar: RCV000144596) which was removed to avoid conflation with monogenic risk.

PRS calculation was performed using a software developed by Mendelics, evaluating the weighted sum of beta values, in which weights are based on the number of the individual’s alleles containing the variant of the PRS file. The sum is normalized by all beta positive and negative values so the final value can be between zero and one.

### Statistical analyses

PRS values were standardized according to the control values prior to all statistical analyses. The effect size of PRS on breast cancer status was assessed using logistic regression, adjusting for z-scored PCs 1 to 10. AUC for the full dataset evaluation was obtained using the yardstick R package (yardstick.tidymodels.org/) roc_auc function, in the testing data split (25%). In order to find segmentation effect-sizes, individuals were classified into deciles or percentiles based on left-open and right-closed intervals. OR for deciles was calculated by first selecting only the decile analyzed and the median interval (40-60%) individuals as the control section, and binarizing it (0 = belongs to the control interval 40-60%, 1 = belongs to the decile analyzed, for example, 10%); and performing a logistic regression analysis on the binarized decile information with correction for PCs 1 to 10. A similar approach was conducted for calculating the OR on percentiles for comparison with Mavaddat’s^8^ PRS validation. All comparisons with original studies were made with the values for the testing sets. For each ancestry proportion group, AUC was estimated using 10-fold cross-validation with the R package *caret*^20^. OR and CI for genes *BRCA1*, *BRCA2*, *PALB2*, *TP53*, *ATM* and *CHEK2*, and for the variant R337H from *TP53* were obtained using *epitools* R package^21^. All statistical tests performed were two-tailed. All analyses were performed in R version 4.4.2.

## Results

### Case-control sample selection and characteristics

After removal of 211 subjects with a first-degree relationship, 73 with missing files necessary for imputation, and 122 with a low-quality imputation, a total of 15,084 subjects remained (**Supplementary Table 1**). Individuals with P/LP variants in BC genes with OR > 5: *BRCA1*, *BRCA2*, *TP53*, *PALB2*, and *PTEN* were removed prior to PRS calculation (n = 629) (**Supplementary Methods**), resulting in a sample consisting of 5,598 women with a BC history, and 8,767 unphenotyped controls (**Table 1**).

**Table 1.** Demographics of cases and controls in BC dataset used for PRS evaluation.

|  |  | **Case** | **Control** | **Total** | **p value** |
| --- | --- | --- | --- | --- | --- |
|  | Total | 5,598 | 8,767 | 14,365 | - |
| **Sex** | F | 5,598 | 4,187 | 9,785 | - |
|  | M | - | 4,580 | 4,580 | - |
| **Age mean**<br>**(SD)** | Total | 49.8 (11.6) | 41.6 (13.3) | 44.8 (13.3) | 0.000 |
|  | F | 49.8 (11.6) | 41.9 (13.7) | 46.4 (13.1) | 0.000 |
|  | M | - | 41.3 (12.9) | 41.3 (12.9) | - |

p-values obtained from two-tailed t-tests.

The ancestry composition of our admixed cohort, obtained using admixture^22^ (**Supplementary Methods**), is shown in **Figure 1**.The majority of individuals have EUR as their greatest ancestry proportion (median 84%, SD 18%). Significant fractions of AFR (median 6%, SD 12%) and AMR (median 8%, SD 7%) ancestries are observed, along with a variety of EUR proportions. A small proportion of EAS ancestry is also observed (median < 1%, SD 12%), primarily consisting of 214 individuals with over 70% of this ancestry.

**Figure 1.**
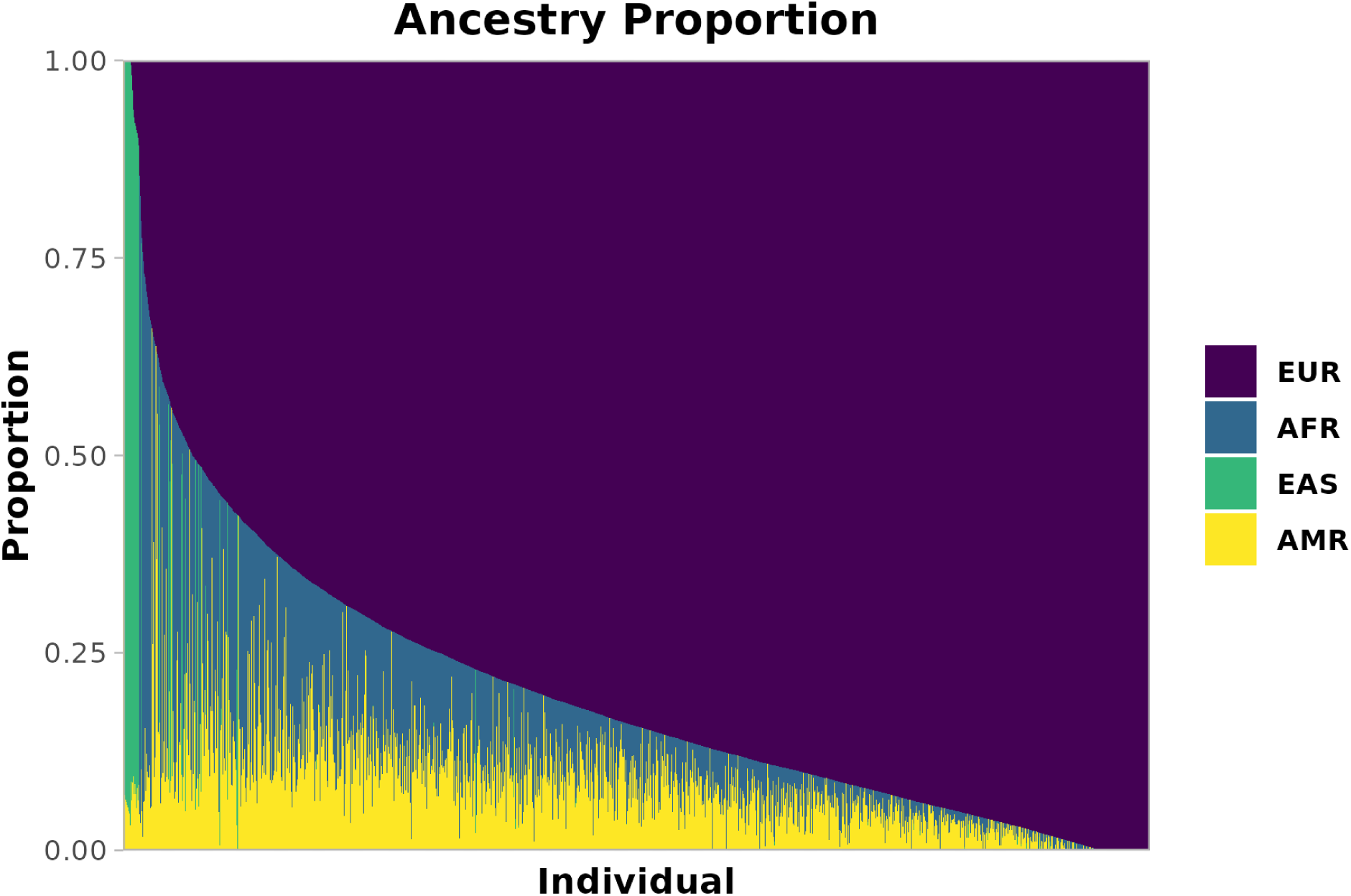
Ancestry composition of our Brazilian cohort. Estimated ancestries are shown as proportions per individual. Each thin bar represents one individual and their ancestry proportion. Europe (EUR) in purple, Africa (AFR) in blue, East Asia (EAS) in green and America (AMR) in yellow.

### Effect sizes of four different PRSs in the Brazilian population

Four PRS files from three studies were selected for initial effect size investigation in our cohort: Broad ^7^, 313 ^8^, 3820 ^8^ and UKBB ^15^ (**Supplementary Table 2**). All PRS files had their variants filtered to address only variants covered by the imputation of our exomes.

PRSs were calculated for the exomes imputed into genomes (details described in the **Methods**) and standardized to improve interpretability. Effects were corrected for the ten first PCs, and the results are reported in **Supplementary Table 3**.

Three of the four PRSs examined had statistically significant associations with breast cancer risk, with the ORs per SD ranging from 1.35 to 1.52 (PRS_Broad_: OR 1.52, 95% CI 1.46 - 1.59, AUC 0.614; PRS_3820_: OR 1.43, 95% CI 1.38 - 1.49, AUC 0.596; PRS_313_: OR 1.35, 95% CI 1.30 - 1.41, AUC 0.583). UKBB PRS was not significantly associated with breast cancer risk in our cohort (p value 0.40). Goodness of fit of the model was greater for PRS_Broad_ (pseudo-R²: 0.062) and PRS_3820_ (pseudo-R²: 0.054).

Since PRS_Broad_, PRS_3820_ and PRS_313_ showed significant results per SD, they were used to split the data into deciles to evaluate BC risk conferred by PRS in each strata. These analyses were also corrected for the first ten PCs. In **Figure 2** we can observe the staircase shape for all of the three PRSs. Especially the bottom and top deciles, the most critical when analyzing PRS data, show statistical significance (p < 0.001) and important effect sizes for all PRSs, ranging from 0.48 to 0.55 in the lowest decile, and 1.73 to 2.13 in the highest (**Supplementary Table 4**). As the most notable result, PRS_Broad_ shows over two-fold increased risk of breast cancer for women in the top decile (90-100%) compared to the middle decile (40-60%).

**Figure 2.**
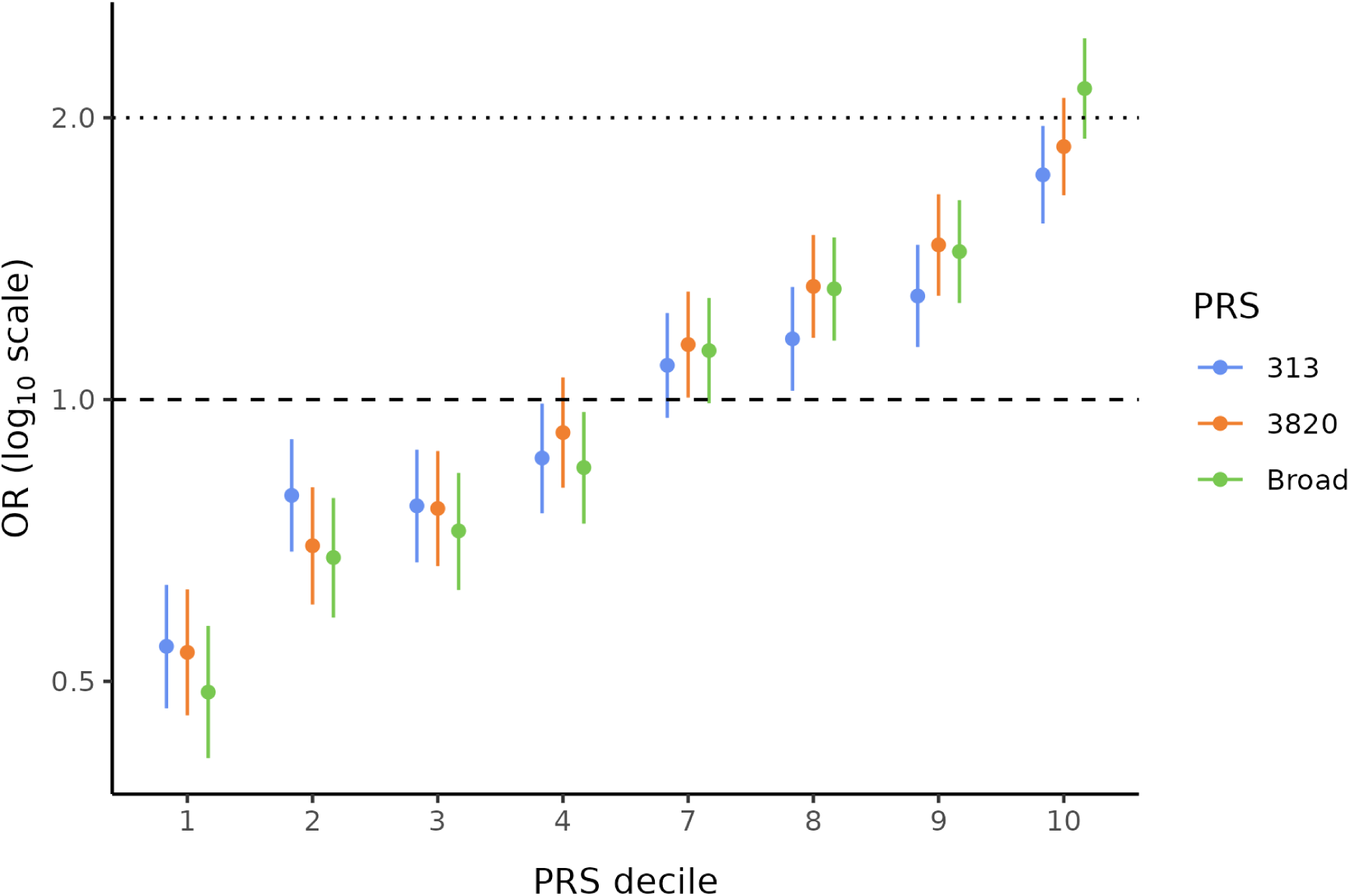
Effect sizes by decile of PRS_313_, PRS_3820_ and PRS_Broad_. Odds Ratios (OR) and Confidence Intervals (CI) are represented for PRS_313_ (blue), PRS_3820_ (orange) and PRS_Broad_ (green). ORs for all PRS deciles were corrected for the first ten PCs. Deciles 5 and 6 were used as references to calculate ORs for the other deciles.

### Comparison of PRSs performances with the original studies

The comparison of PRS metrics of this work with their original studies allows us to evaluate whether the metrics retain their accuracy and reliability in a genetically diverse and admixed population. Concerning OR per SD, all PRSs show a less pronounced value for our cohort compared to the original studies (OR PRS_Broad_: 1.52 vs. 1.56; PRS_3820_: 1.43 vs. 1.66, PRS_313_: 1.35 vs. 1.61). Similarly, the classification ability is less robust in our analysis (AUC Broad: 0.61 vs. 0.69; 3820: 0.60 vs. 0.64, 313: 0.58 vs. 0.63). This is an expected result given that our admixed population presents differences in allele frequencies and linkage disequilibrium, compared to the pure European populations used for both construction and validation of the PRSs.

Considering OR per percentiles, we observed that the PRS_3820_ shows a closer resemblance to Mavaddat’s results than PRS_313_ (**Figure 3**). This is primarily due to the top 1% OR, which shows a stronger effect for PRS_3820_ than for PRS_313_ (OR 2.93 vs. 1.98), aligning more closely with the original PRS_3820_ result (OR 3.95) and deviating further from the original PRS_313_ result (OR 4.04).

**Figure 3.**
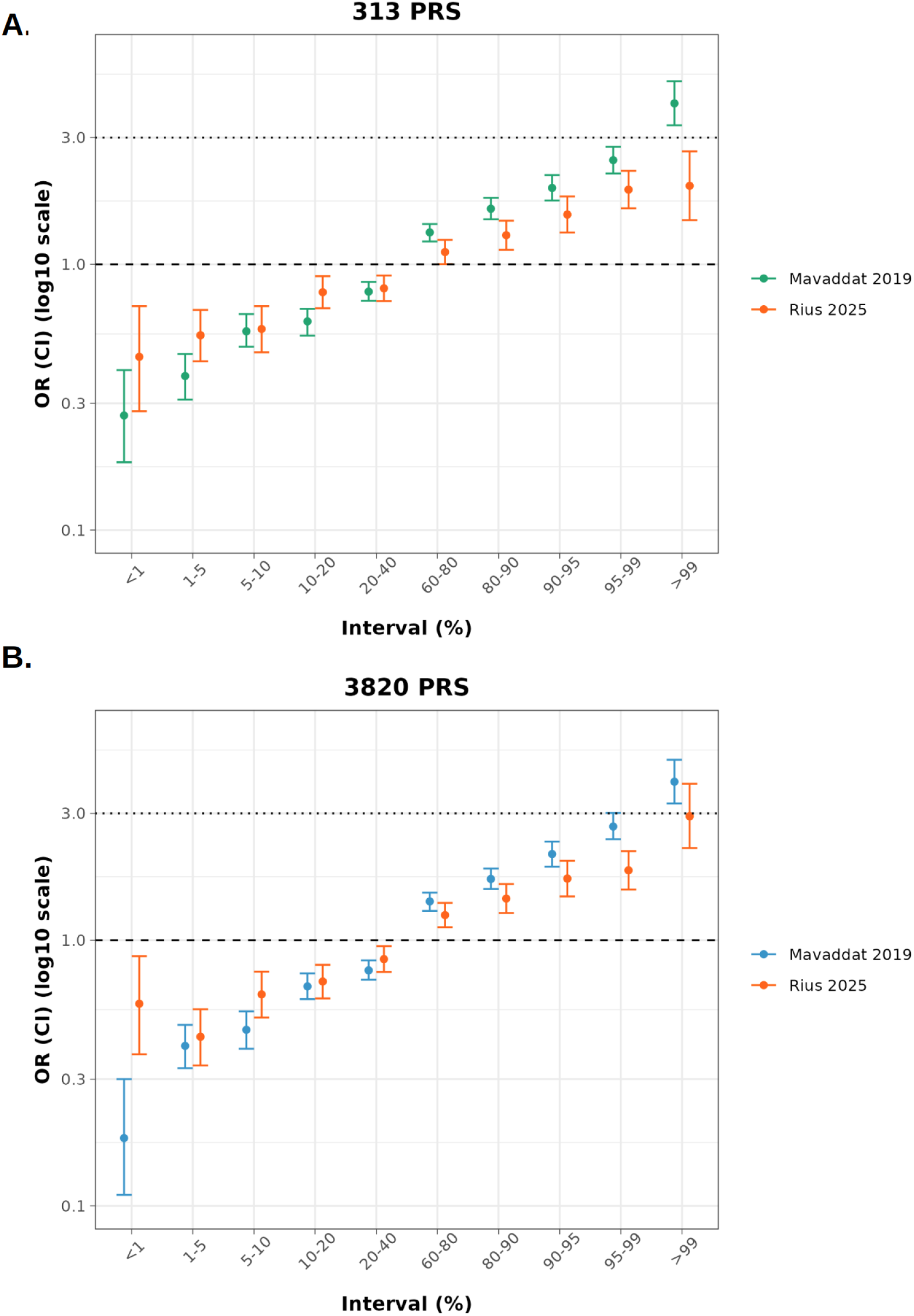
Comparison of PRS_313_ and PRS_3820_ percentile effect sizes between original study and Brazilian cohort. (A) PRS_313_ adjusted to this study (orange), with 283 variants, alongside the original from Mavaddat *et al.* study (green), with 313 variants. (B) PRS_3820_ adapted in this study (orange), with 2,575 variants, alongside the original from Mavaddat *et al.* study (blue), with 3,820 variants.

For PRS_3820_, the lower interval, comprehending the lowest 1% of PRS values, showed a smaller decrease in BC risk compared to the original study. This result is probably related to the small sample size of this section, with only 30 cases and 88 controls available to calculate OR.

### PRS evaluation by ancestry composition

Since our sample contains a great majority of EUR ancestry proportion, we decided to evaluate the PRSs effect sizes in different ancestry compositions (**Figure 4A**, **Supplementary Methods**). All three bins had statistically significant (p < 0.001) ORs above 1.35 per SD for both PRS_313_ and PRS_3820_, showing a positive association of the PRS value with increased BC risk for all ancestry groups (**Figure 4B**). PRS_Broad_ shows significance for both European-related groups (p < 0.001) with the most prominent effect sizes among all PRSs (Non-European majority: OR 1.58, 95% CI 1.34 - 1.88; European majority: OR 1.52, 95% CI 1.47 - 1.58). Nevertheless, PRS_Broad_ did not reach statistical significance for the East Asian majority group (p value = 0.08), indicating that this PRS may not be appropriate for use in the Brazilian admixed population.

**Figure 4.**
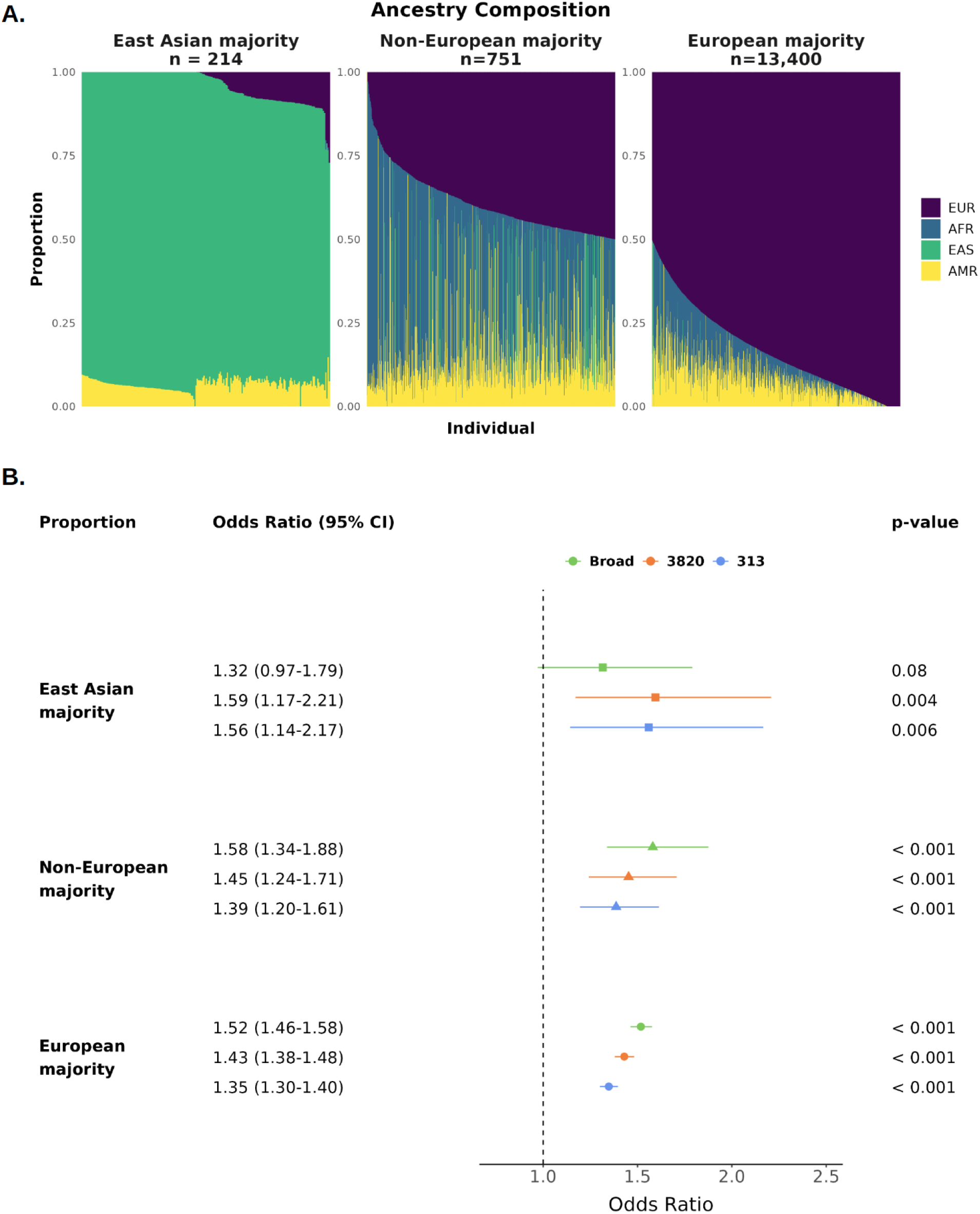
PRSs effect sizes by ancestry proportion. The cohort was split into three groups based on main ancestry: East Asian majority (>50% EAS), Non-European majority and European majority (A) Ancestry composition of each group, with colors representing continental ancestries for each subject: purple for EUR, blue for AFRm green for EAS and yellow for AMR. (B) Breast cancer ORs by PRS_Broad_, PRS_3820_ and PRS_313_ SD for the three ancestry groups. All p-values displayed were corrected for multiple-hypothesis testing using Bonferroni method.

### Correlation of PRS results derived from exomes and from genomes

The subsequent analyses focused on PRS_3820_. A correlation of 0.74 (p value < 2.2e-16) was obtained between BC PRS_3820_ values calculated from exomes followed by imputation and sequenced genomes, showing a concordance between both methods (**Supplementary Figure 1A**). Comparison of the proportion of individuals classified into deciles (**Supplementary Figure 1B**) shows that for the top and bottom deciles there is major concordance (57%) of the respective decile. Furthermore, the individuals classified in different deciles are mostly present in the surrounding deciles, indicating consistent results from both imputed and sequenced data.

### Breast cancer genes and PRS effect size comparison

For the purpose of understanding how the PRS_3820_ effect size compares to known high and moderate risk genes for BC, we have compared OR of the top PRS_3820_ decile (PRS90) with all pathogenic variants located in *TP53*, *BRCA1*, *BRCA2*, *PALB2*, *ATM* and *CHEK2* genes, plus the pathogenic variant R337H of *TP53* gene (**Figure 5**) in this cohort of individuals.

**Figure 5.**
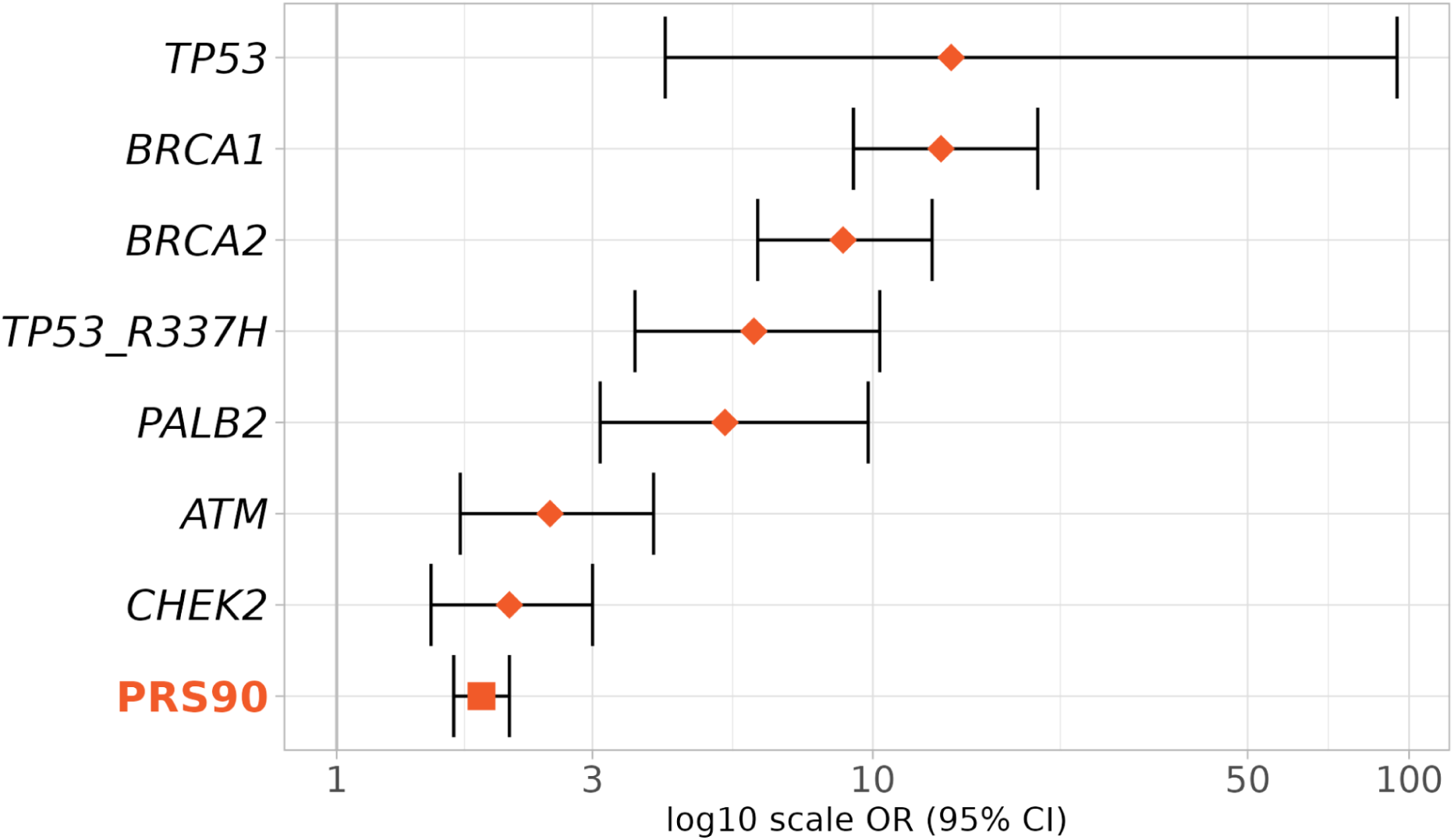
Effect sizes of top decile of PRS and BC genes in BC risk. Effect sizes (OR and 95% CI) were obtained according to the presence of pathogenic variants in the genes *TP53*, *BRCA1*, *BRCA2*, *PALB2*, *ATM* and *CHEK2*, or inclusion in the 90th to 100th percentiles of PRS_3820_ (PRS90).

As expected, *TP53*, *BRCA1* and *BRCA2* present the most extreme BC risks (*TP53* OR: 14, 95% CI: 4.1-95; *BRCA1* OR: 13.4, 95% CI: 9.2-20.3; *BRCA2* OR: 8.8, 95% CI: 6.1-12.9, respectively). PRS90 risk (OR: 1.9, CI: 1.7-2.1) is slightly lower but comparable to the risk conferred by moderate risk BC genes *ATM* (OR: 2.5, CI: 1.7-3.9) and *CHEK2* (OR: 2.1, CI: 1.5-3).

## Discussion

In the present study we have validated two breast cancer PRSs developed from Europeans in the highly admixed Brazilian population. The PRSs adapted from Mavaddat *et al.* study with 283 (PRS_313_) and 2,575 (PRS_3820_) variants^8^ showed statistically significant risk prediction values both per PRS SD and for the top decile compared with the middle deciles (p values < 0.001). PRS_3820_ showed the best performance, with an OR of 1.43 per PRS SD (95% CI: 1.38-1.49) and 1.88 for the top decile (95% CI: 1.66 - 2.12). Furthermore, this PRS showed an OR per SD of 1.43 or above along different ancestry compositions (East Asian majority: OR 1.59, 95% CI 1.17-2.21, p value 0.004; Non-European majority: OR 1.45, 95% CI 1.24-1.71, p value < 0.001; and European majority: OR 1.43, 95% CI 1.38-1.48, p value < 0.001), highlighting its suitability for the diverse Brazilian population.

The PRS with the best performance of this study is based on a previous study from Mavaddat et al. 2019, which developed and validated a PRS with 3,820 variants evaluating invasive BC risk. For all BC subtypes (ER+ and ER-) they found an OR of 1.71 per SD (CI: 1.64 1.79) in the validation set (n = 29,751; cases = 11,428), and OR 1.66 per SD (CI: 1.61 - 1.70) in the prospective set (n = 190,040; cases = 3,215). These values are even greater compared to the widely used 313 PRS (OR: 1.65 per SD; CI: 1.59 - 1.72 in validation set). However, they included a *CHEK2* gene pathogenic variant in the PRS and worked with only invasive BC, which may have led to overestimating their OR values. A study from Liu and colleagues has evaluated another modification of the same PRS with 3,820 variants developed from Mavaddat *et al.* for African, Latin, and European populations^23^. According to the study, the effect size of this PRS in an European sample (n = 33,594) was 1.40 per SD, a result very similar to ours for a Brazilian sample (OR 1.43 per SD; n = 14,365). They deliberately have included women with *in situ* ductal BC as well as women with invasive BC, what they claim to be a reason for OR decline compared to the original study. Our study, however, does not distinguish BC types, therefore we hypothesize that both invasive and *in situ* BC are included, which may be a factor, together with genetic population structure, that decreased the OR compared to the original study.

All of our PRS values were calculated according to a novel methodology: the imputation of exomes. This approach has demonstrated to be very successful for PRS calculation and assessment of BC risk in our study, and could be very interesting for laboratories that already perform exome sequencing as a cost-effective methodology to identify P/LP variants for BC. Multiple studies have compared low-pass genome sequencing with arrays for different applications, such as pharmacogenetics, GWAS and PRS calculation^24,25,26^. The study of Li *et al.*^25^ reported improved accuracy for polygenic risk prediction of imputed low-pass genome compared to array imputation for both coronary artery disease and BC. Despite the slight difference we found between PRS values calculated from sequenced genomes and exomes with imputation (rho: 0.74), decile classification showed satisfactory concordance between both methods for the majority of results in the extreme deciles (1 and 10th), which are the most important to define decreased or increased risk. Unfortunately, it was not possible to assess the predictive power of PRS values calculated from genomes of BC patients due to unavailability of paired exome and genome data.

Among familial BC cases, approximately 25% have a P/LP germline variant reported^27^. In the Brazilian population, a robust study with 1,663 breast cancer patients detected 20.1% of P/LP germline variants using multigene panel testing^4,5^. A 2017 study reported that 18% of the hereditary BC can be explained by a polygenic effect of variants discovered in a GWAS^28^. Therefore, employing this PRS in the clinical practice might bring an elucidation to BC Brazilian families without high or moderate-effect germline variants detected. Moreover, women without prior knowledge of their familial BC condition, or even those with a high PRS risk by chance, will have the possibility to be informed of their results and share them with their physicians to adopt preventive actions accordingly to their risk strata, such as intensifying surveillance adding breast magnetic resonance imaging to mammography screening^29^.

In conclusion, we have validated both PRS_3820_ and PRS_313_ in the Brazilian population, demonstrating their potential utility for breast cancer polygenic risk assessment. Notably, PRS_3820_ exhibited a greater effect size, with the top decile presenting a risk comparable to moderate-risk monogenic variants for BC. Future studies will be required to evaluate the combination of PRS with P/LP variants and clinical factors in order to deliver more informative results to patients, thus physicians can recommend prevention strategies based on their combined polygenic and monogenic BC risk.

## Ethics Statement

This work was approved by the Ethics Committee of University of São Paulo - Faculdade de Medicina under the CAAE number 70112423.3.0000.0068.

## Supporting information

313 PRS

3820 PRS

Broad PRS

UKBB PRS

## Acknowledgements

We thank all individuals once sequenced in Mendelics laboratory who have consented to participate in this research. We also thank all UKBB participants for their contribution to the PRS hereby analyzed, and all authors from previous studies on BC PRSs in which we based our validation (Khera *et al.* 2018 and Mavaddat *et al.* 2019).

## Funding

Maria Aparecida Azevedo Koike Folgueira received research support from Conselho Nacional de Desenvolvimento Científico e Tecnológico, Brazil (CNPq—308052/2022-6).

## Data Availability

All variants and betas which compose the four evaluated PRSs are available as Supplementary Information. Individual cases and controls data are not publicly available due to the confidentiality consentment agreement signed by all included in the study.

## Competing Interests

Flávia Eichemberger Rius, Danilo Viana, Júlia Salomão, Laila Gallo, Renata Freitas, Cláudia Bertolacini, Lucas Taniguti, Danilo Imparato, Flávia Antunes, Gabriel Sousa, Renan Achjian, Eric Fukuyama, Cleandra Gregório, Iuri Ventura, Juliana Gomes, Nathália Taniguti, and David Schlesinger are currently employed by Mendelics, or were employed at the time of the study.

Rodrigo Guindalini acted as a consultant for AstraZeneca, Janssen Oncology, Roche/Genentech and Igenomix; received speaker honoraria from AstraZeneca, Bristol Myers Squibb, GlaxoSmithKline, Merck Sharpe & Dohme Brasil, Novartis, and Roche outside the submitted work; and has equity in Mendelics Análise Genômica.

Olufunmilayo I. Olopade is co-founder at CancerIQ; serves as scientific advisor at Tempus; and has received research funding from Color Genomics and Roche/Genentech.

José Eduardo Krieger, Yonglan Zheng, Dezheng Huo, Simone Maistro and Maria Aparecida Koike declare no competing interests.

## Author Contributions

### Generated Main Data

Flávia Eichemberger Rius, Danilo Viana, Júlia Salomão, Laila Gallo, Renata Freitas, Cláudia Bertolacini, Lucas Taniguti, Danilo Imparato, Flávia Antunes, Gabriel Sousa, Renan Achjian, Eric Fukuyama, David Schlesinger.

### Analyzed Data

Flávia Eichemberger Rius, Rodrigo Guindalini, Danilo Viana, Lucas Taniguti, Danilo Imparato, Flávia Antunes, Gabriel Sousa, Renan Achjian, Eric Fukuyama, Yonglan Zheng, Dezheng Huo, Olufunmilayo I. Olopade, Maria Aparecida Koike, David Schlesinger.

### Other Contributions

Cleandra Gregório, Iuri Ventura, Juliana Gomes, Nathália Taniguti, Simone Maistro, José Eduardo Krieger.

## Notes

### Author Declarations

This work was approved by the Ethics Committee Comissão para análise de projeto de pesquisa of Hospital das Clínicas da FMUSP - CAPPesq under the CAAE number 70112423.3.0000.0068

### Summary of Updates

Results and figures updated after correction of PRS 313 variant set; results and figures updated after removal of 122 individuals with a low quality of imputation; all supplementary information related to the previous topics updated; formatting changed.

