## Supplementary Information for "A Breast Cancer Polygenic Risk Score Validation in 15,490 Brazilians using Exome Sequencing"

#### Supplementary Methods

##### Relatedness calculation and data filtering

Relatedness of individuals was obtained from the exomes using somalier software[1], following the standard protocol required for a VCF file (https://github.com/brentp/somalier#readme). Concerning related individuals removal, if two individuals had a first-degree relationship, one of them was randomly selected to be included in the dataset. However, if individuals had two or more first-degree relationships, all related individuals were excluded from the dataset. This process resulted in a total of 211 removals. Furthermore, 73 individuals were removed from the sample due to unavailability of files necessary for genome imputation, and additional 122 individuals were removed due to low-quality imputation.

PRS analyses were performed after filtering out cases and controls with pathogenic or likely-pathogenic (P/LP) variants in high penetrance BC genes with OR > 5: *BRCA1*, *BRCA2*, *TP53*, *PALB2*, and *PTEN*.

##### Ancestry Evaluation

Admixture[2] was used to extract continental ancestries from all non-related and data completed exomes. The analysis was supervised by the 1KGP samples, after removal of South Asian, Oceania, and admixed Americans from the GRCh38 1KGP release of 2017. South Asian and Oceania ancestries were removed because they are not a significant part of Brazilian ancestral composition. Latin American admixed populations (Colombian, Peruvian, Puerto Rican, and Mexican) were removed to avoid confounding with the native americans belonging to the same population label. Continents evaluated were: Africa - AFR, America - AMR, East Asia - EAS, and Europe - EUR. Ancestry results were further used for splitting individuals into groups according to their ancestry composition, to analyze the effect size of PRS on each group. The groups created were: East Asian majority (>50% EAS, n = 217), Non-European majority (0-50% EUR, n = 760) and European majority (51-100% EUR, n = 13,510).

**Genetic Principal Component Analyses (PCA) Assessment**

PCA was calculated for exomes from a projection in 1KGP[3] and Human Genome Diversity Project (HGDP)[4] samples. Only variants with MAF > 1% and that could have been directly genotyped using WES were included for the PCA analysis in 1KGP and HGDP samples using plink2[5]. Exomes were converted to plink bfile format (bed, bim, and fam files) and had duplicated variants removed. PCA projection for 10 PCs was calculated using plink2 –score method, with allele frequencies from the breast cancer cohort.

**Paired Imputed and Sequenced Genomes Analysis**

Exome-imputed variants and directly sequenced variants from WGS were compared using 3,119 samples from an independent Brazilian population dataset (http://elsabrasil.org/) that had both WES and WGS available. The WES were sequenced and imputed also using the same method previously described. Adapted BC PRS-3820 from Mavaddat *et al.* study was calculated for both exomes with imputation and genomes, and their Spearman correlation was calculated using R software function *cor.test*.

### Supplementary Tables

**Supplementary Table 1**. Demographics of cases and controls and BC characteristics.

| | | Case | Control | Total | p.value |
|---|---|---|---|---|---|
| | Total | 6,206 | 8,878 | 15,084 | - |
| **Sex** | F | 6,206 | 4,241 | 10,447 | - |
| | M | - | 4,637 | 4,637 | - |
| **Age** | Total | 49.5 (11.7) | 41.6 (13.3) | 44.9 (13.2) | 0.000 |
| | F | 49.5 (11.7) | 42 (13.7) | 46.5 (13.1) | 0.000 |
| | M | - | 41.3 (12.9) | 41.3 (12.9) | - |
| **P/LP variants*** | No | 5,598 | 8,767 | 14,365 | - |
| | Yes | 608 | 111 | 719 | - |
| **Subtype**** | HER2+ | 162 | - | 162 | - |
| | HR+ | 306 | - | 306 | - |
| | Triple Negative | 1,025 | - | 1,025 | - |
| | No information | 4,713 | - | 4,713 | - |
| **Multiple Breast Tumors** | No | 5,729 | - | 5,729 | - |
| | Yes | 477 | - | 477 | - |

* Pathogenic variants on the genes *BRCA1*, *BRCA2*, *TP53*, *PALB2*, and *PTEN* were evaluated to classify individuals in this category.

** For multiple tumors with information we have selected the subtype of the most recent tumor.

**Supplementary Table 2**. Number of individuals and variants of the four PRSs evaluated in this study.

| Study | PRS name | PGS Catalog ID | Total in discovery sample | Cases | Controls | Original variants |
|---|---|---|---|---|---|---|
| Khera et. al, 2018 | Broad | PGS000015 | 228,951 | 122,977 | 105,974 | 5,218 |
| Mavaddat et. al, 2019 | 313 | PGS000004 | 169,092 | 94,075 | 75,017 | 313 |
| Mavaddat et. al, 2019 | 3820 | PGS000007 | 169,092 | 94,075 | 75,017 | 3,820 |
| UKBB (phenotype 20001_1002) | UKBB | - | 194,153 | 7,968 | 186,185 | 7,538 |

* Number of variants after adjustment for imputation coverage.

**Supplementary Table 3**. Metrics for All PRSs Evaluated.

| PRS | OR | p value | Lower 2.5% CI | Upper 97.5% CI | AUC | Nagelkerke pseudo-$R^2$ |
|---|---|---|---|---|---|---|
| Broad | 1.52 | 6.10E-82 | 1.46 | 1.59 | 0.614 | 0.062 |
| 3820 | 1.43 | 1.02E-68 | 1.38 | 1.49 | 0.596 | 0.054 |
| 313 | 1.35 | 1.64E-49 | 1.3 | 1.41 | 0.583 | 0.042 |
| UKBB | 1.02 | 0.40 | 0.98 | 1.06 | 0.545 | 0.014 |

* AUC: Area under the receiving operated curve, evaluating the performance of PRS only, without covariates, on BC classification.

** Variance explained by PRS: calculated as a difference of Nagelkerke's pseudo-$R^2$ value obtained in a model with and without the evaluated PRS.

**Supplementary Table 4**. PRS Broad, PRS 3820 and PRS 313 decile Odds Ratios and Confidence Intervals.

| Broad | | | | |
|---|---|---|---|---|
| **ORs** | **lower 95% CI** | **upper 95% CI** | **pval** | **decile** |
| 0.48 | 0.41 | 0.57 | 1.90E-18 | 10% |
| 0.69 | 0.59 | 0.79 | 4.39E-07 | 20% |
| 0.71 | 0.61 | 0.82 | 2.64E-06 | 30% |
| 0.83 | 0.72 | 0.95 | 0.0072 | 40% |
| 1.12 | 0.98 | 1.27 | 0.1018 | 70% |
| 1.29 | 1.14 | 1.47 | 0.0001 | 80% |
| 1.44 | 1.26 | 1.63 | 2.46E-08 | 90% |
| 2.13 | 1.88 | 2.41 | 3.97E-33 | 100% |

| 3820 | | | | |
|---|---|---|---|---|
| **ORs** | **lower 95% CI** | **upper 95% CI** | **pval** | **decile** |
| 0.54 | 0.46 | 0.63 | 1.37E-14 | 10% |
| 0.70 | 0.60 | 0.81 | 1.16E-06 | 20% |
| 0.77 | 0.67 | 0.89 | 2.66E-04 | 30% |
| 0.93 | 0.81 | 1.07 | 0.2990 | 40% |
| 1.14 | 1.00 | 1.30 | 0.0514 | 70% |
| 1.34 | 1.18 | 1.52 | 0.0000 | 80% |
| 1.44 | 1.27 | 1.63 | 1.44E-08 | 90% |
| 1.88 | 1.66 | 2.12 | 1.19E-24 | 100% |

| 313 | | | | |
|---|---|---|---|---|
| **ORs** | **lower 95% CI** | **upper 95% CI** | **pval** | **decile** |
| 0.55 | 0.47 | 0.64 | 7.93E-15 | 10% |
| 0.79 | 0.68 | 0.90 | 6.81E-04 | 20% |
| 0.77 | 0.67 | 0.88 | 1.66E-04 | 30% |
| 0.86 | 0.75 | 0.99 | 0.0330 | 40% |
| 1.09 | 0.96 | 1.24 | 0.2072 | 70% |
| 1.14 | 1.00 | 1.30 | 0.0429 | 80% |
| 1.29 | 1.14 | 1.46 | 8.62E-05 | 90% |
| 1.73 | 1.54 | 1.95 | 3.56E-19 | 100% |

### Supplementary Figures

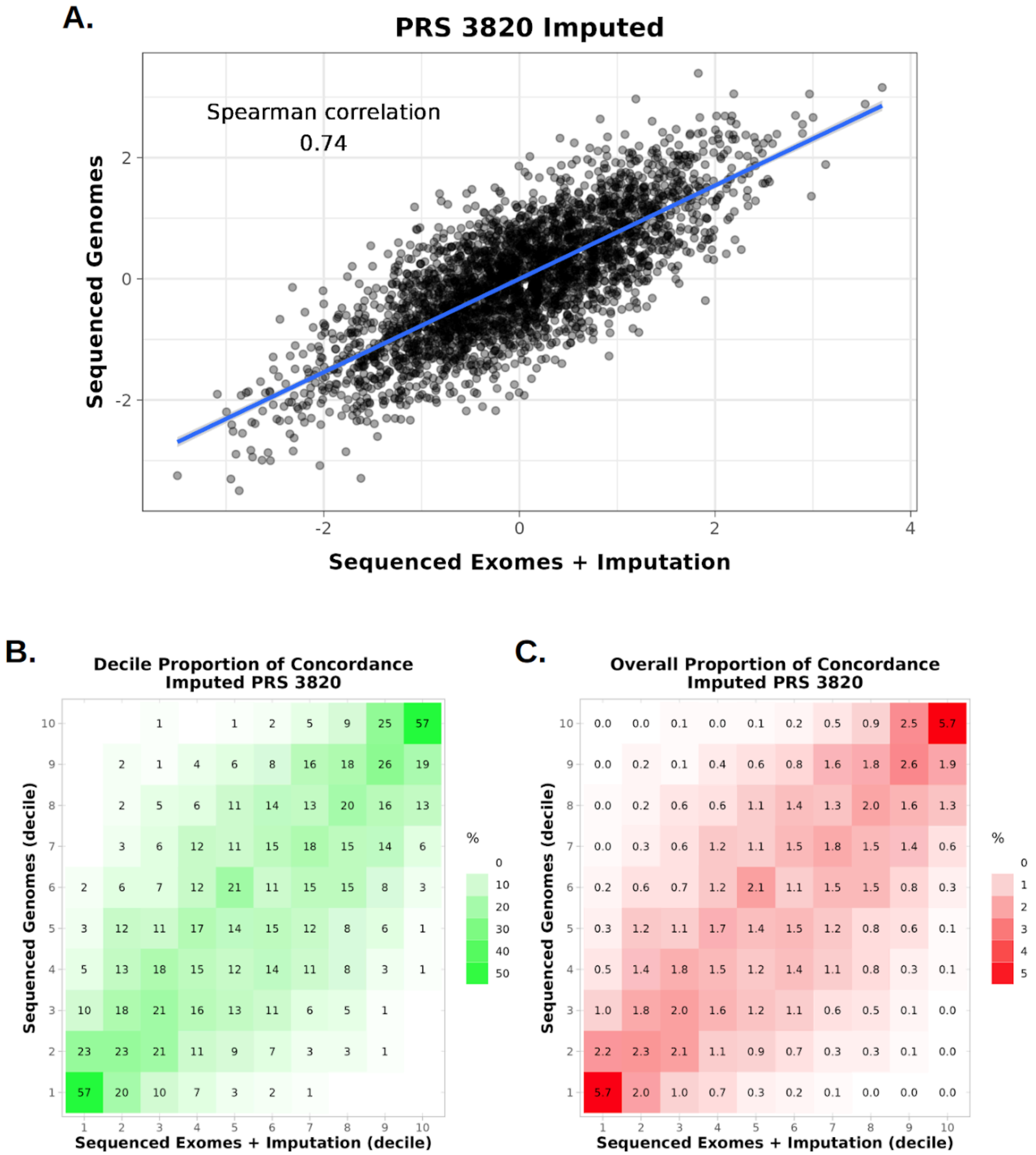

**Supplementary Figure 1. Correlation of $PRS_{3820}$ values for exomes with imputation and genomes.** (A) Correlation between PRS values for exomes with imputation and sequenced genomes. Spearman correlation rho value is shown in the plot area. (B) Heatmap showing the percentage of samples classified in a determined combination of deciles between exomes with imputation (x axis) and genomes (y axis). Proportion expressed in percentage per decile. (C) Heatmap showing the percentage of samples classified in a combination of deciles between exomes with imputation (x axis) and genomes (y axis). Proportion expressed in overall percentage.
